# Validity of the German version of the Stay Independent Questionnaire applied by telephone interview: A diagnostic accuracy study

**DOI:** 10.1101/2025.02.09.25321960

**Authors:** Ulrich Thiem, Ingeborg Schlumbohm, Stefan Golgert, Ulrike Dapp, Saskia Otte

## Abstract

**Background:** Mobility limitations are among the most common functional problems in older people. Repeated falls can lead to injuries and fractures, trigger or intensify fear of falling, and contribute to subsequent functional decline and loss of independence. Various questionnaires have been developed, both nationally and internationally, to identify older people at increased risk of falling. Data for evaluation against standard tests from the geriatric mobility assessment are scarce.

**Methods:** In a German project evaluating home emergency call systems, the Stay Independent Questionnaire (SIQ) from the American prevention program STEADI (Stopping Elderly Accidents, Deaths, and Injuries) was used for the identification of community-dwelling seniors aged 70 and older at risk of falling. The original questionnaire was translated by professional translators using the typical forward and backward translation process, and a final version was established after discussion involving a bilingual scientist. The diagnostic performance of the questionnaire (diagnostic test) was tested against the mobility assessment Short Physical Performance Battery (SPPB, gold standard). To describe the test performance, typical statistical measures are used, i.e., sensitivity, specificity, positive and negative predictive values, and positive and negative likelihood ratios, each with the corresponding 95% confidence interval (95% CI).

**Results:** Data from a total of 190 participants (143 women, 75.3%; average age 80.5 years ± 5.5 years standard deviation) were included in the analysis. According to existing comorbidities and functional abilities, between 30% and 40% suffered of advanced comorbidity and/or functional impairment. The questionnaire identified 148 individuals (77.9%) as at risk of falling. According to SPPB, 81 participants had an objectively measurable impairment of standing and walking balance. The test performance measures for SIQ as a diagnostic test are: sensitivity 95.1% 95% CI [ 88.0% ; 98.7%], specificity 34.9% [26.0% ; 44.6%], positive and negative predictive value 52.4% [44.0% ; 60.6%] and 90.5% [77.4% ; 97.3%], respectively, and positive and negative likelihood ratio 1.46 [1.26 ; 1.69] and 0.14 [0.05 ; 0.38]. In receiver-operating characteristic (ROC) analysis, the unadjusted area under the curve for SIQ was 65.0% [57.3% ; 72.7%], after adjustment for sex and age 71.0% [63.8% ; 78.2%].

**Conclusions:** The German version of the Stay Independent Questionnaire is capable of identifying community-dwelling seniors aged 70 and older with unimpaired standing and walking balance. The high sensitivity of the test allows excluding test-negative individuals from further investigation. A limitation of the questionnaire is the high proportion of false positives, resulting from the low specificity of the test. Scientific evaluation will show to what extent the use of the questionnaire may improve the identification and medical care of community-dwelling seniors at risk of falling in terms of fall and fall consequence prevention.

## Background

Falls, as an expression of mobility limitations, are a common and serious functional problem in older individuals and geriatric patients. It is estimated that at least one fifth of people aged over 65 years fall at least once a year (1). The frequency varies in different populations and is highest among the very old and individuals in nursing care facilities (1,2). Repeated falls lead to impaired quality of life through pain and injuries (3,4). They can trigger or worsen fear of falling and may contribute to further functional decline and loss of independence (5). Feared consequences of falls include fractures, particularly proximal femoral fractures, but also fractures and injuries of other locations. In Germany, falls directly cause about 10,000 deaths annually (6). Among adults, higher age groups, especially those aged 80 years and older, and women in particular are affected (6,7).

The burden on the healthcare system due to costs associated with falls is considerable. In the USA, healthcare costs for managing fatal fall-related injuries were over $600 million, and costs for non-fatal fall injuries exceeded $30 billion for the years 2012 and 2015. Between 2012 and 2015, an increase of expenditures of about 3.5% was noted, with a projected upward trend in subsequent years due to increasing fall rates, especially among older age groups (8,9). Comparable health economic consequences and trends are reported for China (10). One reason for the financial burden is the repeated utilization of medical services by individuals after one or more fall events (11).

As a low-threshold approach to identify community-dwelling seniors at increased risk of falling, various self-report questionnaires have been developed nationally and internationally. Examples are: Geriatric Postal Screening Survey (GPSS, Alessi, 2003) (12); Fall Risk Check (Sturzrisiko-Check, SRC, Anders, 2006) (13); Falls Risk of Older People in the Community (FROP-COM, Russell, 2008) (14); Falls Risk Questionnaire (FRQ, Rubenstein, 2011) (15); models by Hirase (16) and Gadkaree (17); and the online questionnaire by Obrist (18). The mentioned instruments vary considerably. The Fall Risk Check, the only German questionnaire, covers 13 areas, each with between two and seven individual statements to be checked and made (13,19). The online questionnaire by Obrist, the only other German-language tool, developed in Switzerland, consists of 29 predictors of fall risk with a total of 36 questions (18). A very short model by Gadkaree uses only five items – age, sex, ethnicity, and falls and balance problems by self-report. However, the test performance of this model is rather limited (17). For most tools, the extent of evaluation is limited. Often, only one publication exists that describes the development of the instrument.

The prevention program STEADI (STopping Elderly Accidents, Deaths, and Injury) initiated by the American Center of Disease Control and Prevention (CDC) (20) uses a variant of the Fall Risk Questionnaire (FRQ) developed by Rubenstein (15), called Stay Independent Questionnaire (SIQ). The questionnaire includes twelve simple questions that cover areas such as the number of falls in the past year, fear of falling, sensory impairments, dizziness, medications that may increase falls risk, tendency towards depression, and more. Although the questionnaire itself was not extensively validated, several studies evaluated the STEADI program as a whole and rate it positively (21–24). According to a recently published implementation study, the STEADI program is effective in preventing falls and their consequences, and is also cost-effective (25).

For the recruitment of community-dwelling seniors with increased falls risk for a randomised controlled trial (RCT) investigating home emergency call systems, we needed an easy to use questionnaire suitable for use in a telephone interview. Our choice fell on SIQ, and we decided to translate it into German and validate it against a standard mobility assessment.

## Methods

### a) Study Hypothesis

A study hypothesis to be statistically tested was not formulated. Our hypothesis was that the translated version of SIQ is able to validly identify community-dwelling seniors aged 70 and over with unimpaired standing and walking balance. Sufficient validity was assumed for SIQ as a diagnostic test demonstrating a sensitivity of at least 75% or higher. For diagnostic tests, a high sensitivity is required to exclude unimpaired (healthy) individuals (26).

### b) Recruitment of Study Participants

Participants for this diagnostic study were recruited as part of the recruitment for the main study, a German RCT investigating different home emergency call systems (INES, Intelligentes NotfallErkennungsSystem). Details of the main study will be published elsewhere. In brief, participating health insurance companies contacted potentially eligible insured persons in three federal states in Germany – Hamburg, Bavaria, North Rhine Westphalia – and invited them to participate in the RCT. Criteria for contacting included age ≥ 70 years, presumably living alone, and written consent for further contact. Initially, a selection based on ICD-10 codes (International Classification of Diseases, 10th edition, German Modification) (27) coded in the insurers documents was made to increase the number of individuals with diseases increasing falls risk. The ICD codes in use were: G2x – Parkinson’s syndromes; G8x – paresis; H8x – vestibular function disorders / vertigo; I63.x – stroke; R26.x – mobility disorders; R29.6 – fall tendency, U5x – functional impairments. Recruitment began at July 17^th^, 2023. From mid-October 2023 (10/19/2023), insured persons irrespective of specified ICD codes were contacted and invited to participate. Recruitment ended at March 7^th^, 2024, with the scheduled appointment of the last participant.

Consenting individuals were called by a call centre and screened for eligibility to participate. The main criterion for eligibility was an increased fall risk identified by SIQ. Individuals at risk were invited to participate in the RCT, and relevant data were transferred to cooperating providers of home emergency call systems for further procedures.

Regardless of the SIQ results, insured persons residing in Hamburg and contacted by the call centre were additionally offered participation in this sub-study. Thus, insured individuals aged ≥ 70 years residing in Hamburg and living alone were included in the SIQ validation study, regardless of their fall risk assessment via SIQ. For a period of five weeks, from early August to mid-September 2023, recruitment by the call centre had to be suspended to handle the initially large number of people willing to participate. Otherwise, all consenting individuals were recruited consecutively. No compensation was provided for participation in the study. Since potentially mobility-impaired individuals were to be included and examined, reimbursement of travel expenses was offered. About a third of the participants took advantage of this by getting travel or taxi costs reimbursed. As an additional incentive, a written summary and evaluation of the findings was sent to the participants, if desired.

The study personnel who conducted data collection for this study remained blinded to the data collection of the call centre and, in particular, to the information from SIQ. Due to the sequence of recruitment, a randomised administration of SIQ and SPPB was not possible.

### c) German Translation of the Stay Independent Questionnaire

The original version of the questionnaire was developed and published by L. Rubenstein et al. in 2011 (15). Until study planning, no validation against subsequent, incident falls within a cohort study was reported, nor testing against a gold standard, e.g., from geriatric assessment. The CDC incorporated the questionnaire into the STEADI prevention program in 2013 (20,28). Since then, it has been recommended as a screening tool for assessing fall risk in seniors in the USA.

The CDC’s questionnaire version was translated into German and back-translated by professional translators, with the second translator being blinded to the original version. The back-translation and the original version were compared, and the translation was finalised by the study group with the help of a bilingual scientist, experienced in geriatric medicine. The German questionnaire was then pre-tested with several seniors visiting a geriatric day clinic nearby.

The call centre used the translated version of SIQ for use in the telephone interviews. The staff at the call centre were trained in its application during an on-site visit by study personnel from Hamburg. The background and objective of this study were presented, special features of the questionnaire were discussed, and potential problems in application debated. During the recruitment, study personnel were available to the call centre for questions and further discussion.

### d) Test administration

The German translation of SIQ was used by the call centre to determine potential fall risk in potentially eligible seniors. The CDC’s evaluation scheme for SIQ was used, which classifies all individuals as at risk if they answer four or more of the twelve questions in SIQ positively, i.e., indicating a fall risk (SIQ total score ≥ 4 points) (29). Regardless of the total score, all respondents reporting a fall event during the past year are classified as at risk. This corresponds to a weighting of the fall question (yes = 4 pts.) compared to the other eleven questions of SIQ (29). The application of the questionnaire at the call centre occurred without knowledge of further clinical information about the participants. Since the investigation in this validation study occurred subsequently, there was no information from the subproject about participants at the call centre.

While only fall-risk seniors were considered for recruitment for the RCT, both individuals with and without fall risk according to SIQ could be and were recruited for this validation study. Due to the eligibility criteria of the health insurance companies when contacting potential participants for the INES main study, overall, more seniors with fall risk than without were found via the recruitment of the call centre.

As a gold standard for verifying fall risk, the test battery Short Physical Performance Battery (SPPB) from the geriatric mobility assessment was chosen. This test examines standing and walking balance in capable individuals in three different parts. In the standing part, the ability for parallel, semi-tandem, and tandem stand for 10 seconds each is tested. In the walking part, walking speed over a distance of four meters is measured. The third part is the performance of the modified Chair Rise Test (CRT), where the time is measured that participants need to rise up from a chair seat five times without using arms or hands for support (5-CRT). Participants were considered test-positive (pathological) if they scored less than ten points (out of a possible twelve points) on the SPPB total score (30).

Apart from few exceptions, the mobility assessment was conducted by a single female physiotherapist, experienced in geriatric medicine. In cases of questions about the execution, special characteristics of participants, or questions about criteria evaluation, a consensus was reached in the study group after discussion. A medical questionnaire part was separated from the assessment part. The medical part, including participant information and written, informed consent, was conducted by study physicians from the Department of Geriatrics. The study physicians were introduced to the study and the execution of the medical questionnaire part by an experienced specialist and senior geriatrician. In case of uncertainties or questions about the evaluation of individual information in the medical part, consultations occurred between the involved physicians and the study team. Results of the medical part were partially available before the mobility assessment was conducted, but they were not disclosed to the physiotherapist. Vice versa, study physicians were not informed about the results of the mobility assessment.

The World Fall Guidelines recommend an algorithm for determining fall risk in seniors (1). A distinction is made between high risk individuals, individuals with no or low risk, and an intermediate group. According to the World Fall Guidelines, individuals who have not experienced a fall in the previous year and who have none or only a few known fall-promoting factors are categorized as being not at risk. Individuals are considered high risk if they have suffered two or more falls during the past year, needed medical attention due to a fall-related injury, sustained a fracture or other fall-related injury, were unable to get up independently for at least one hour after a fall, and individuals with confirmed frailty and those with loss of consciousness at the event or suspected syncope. For the intermediate fall risk group, not fulfilling low or high-risk criteria, the guidelines recommend conducting walking speed as a standard assessment for further decision-making, with a threshold value of 0.8 meters per second. No separate mobility assessment is recommended for the other risk groups (1).

Walking speed as a gold standard criterion was rejected by the research group, and instead SPPB was chosen. One reason is that the test quality of SIQ was to be captured for all groups of older adults, not just for the group with intermediate risk. In this regard, the guideline recommendation is not suitable for the purposes of our study. Secondly, walking speed is included in the SPPB as a walking part, allowing to make use of walking speed in secondary analyses. Thirdly, clinically different and important information for communication with affected individuals can be derived from the SPPB’s three examination parts: standing balance, walking speed, and functional leg strength (5-CRT). Other mobility tests, e.g., walking speed, the Timed Up & Go Test, or others, usually only allow statements about general walking ability or a single functional component of walking, e.g., sole speed. Finally, experience with using the SPPB in our geriatric centre was a further argument for using it.

### e) Additional data collection

Additional data was collected to capture important functional areas and additional risk factors for mobility impairment or falls. Apart from a general assessment of functional abilities by the Longitudinal Urban Cohort Ageing Study Functional Ability Index (LUCAS FI) (41), we considered the functional areas of mobility, emotion, and cognition, as well as topics such as comorbidity burden, pain, sarcopenia, and self-assessed quality of life. An overview of the assessment and examination instruments used can be found as a supplement (table S1).

### f) Sample size calculation and statistical analysis

The sample size was calculated according to Hajian-Tilaki (46) for studies on diagnostic tests. It was assumed that the sensitivity of SIQ as a diagnostic test is 75% (Sn = 0.75). High sensitivity is required for tests intended for screening, as tests with high sensitivity are suitable for excluding healthy or unaffected individuals (26). The lower bound of the 95% confidence interval (95% CI) for sensitivity should not fall below 65% (95% CI ≤ 10%). Type I error was set at 5% (α = 0.05). The prevalence (relative frequency) of an increased fall risk was assumed to be 60% (prev = 0.6) for the entire sample. Under these assumptions and using formula 6.6 from the above-mentioned publication (46), a sample size of 180 individuals was calculated. An increase in the sample size by 5% to 189 individuals was planned to compensate for subsequent exclusion of cases from the analysis, e.g., due to missing values in the main variables (SIQ or SPPB), subsequent identification of exclusion criteria, withdrawal of participants from the study, or other reasons.

The test quality of SIQ as a diagnostic test is presented with typical statistical measures, i.e., sensitivity (Sn), specificity (Sp), positive and negative predictive value (PPV / NPV), and positive and negative likelihood ratios (LR+, LR-), each with the corresponding 95% CI. The calculation of 95% CI for likelihood ratios is performed using the log method (47), for the other test performance measures using the Wilson method (48), and all other values, e.g., proportions, using normal approximation (Wald method) (49).

Characteristics of the study participants are presented descriptively, for categorical variables with absolute and relative frequencies, for continuous variables with mean and standard deviation. Tests on group differences are conducted, where necessary, with X² test for categorical and t-test for continuous, symmetrically distributed variables. Adjustment of test results for age and gender is performed using logistic regression, with the SPPB category ‘pathological’, i.e., SPPB score < 10 points, as the dependent variable, and the explanatory (independent) variables SIQ result, age, and sex. In testing, a type I error of ≤ 5% (α ≤ 0.05) is assumed for statistical significance.

The sample size calculation was conducted using spreadsheet software (Microsoft Excel, Office 2019), the calculation of confidence intervals with the software Confidence Interval Analysis (CIA, T. Bryant, University of Southampton, 1994). For all other statistical analyses, SPSS for Windows was used (IBM, SPSS Statistics, Version 27).

### g) Ethics and reporting

Like every scientific investigation involving human beings, this study was conducted in accordance and compliance with the Declaration of Helsinki in its current version (50). This sub-study was submitted for review to the Ethics Committee of the Hamburg Medical Association as part of the ethics application of the RCT. The Ethics Committee issued a positive vote on June 26, 2023 (registration number 2023-101032-BO-ff). The RCT has been registered in the German Clinical Trials Register (Deutsches Register Klinischer Studien, German Clinical Trials Register DRKS00031408, June 28^th^, 2023). All participants gave written informed consent.

Our report follows the recommendations for the publication of diagnostic tests of the STARD Initiative (STAndards of Reporting Diagnostic Accuracy Studies) (51). The STARD checklist is added as a supplement (table S4).

## Results

### a) Recruitment

An overview of the recruitment is shown in figure 1. The call centre successfully contacted a total of 680 insured individuals residing in Hamburg for the RCT. Contact with 272 individuals occurred outside the recruitment period of this sub-study. All other 408 individuals, regardless of the SIQ results, were offered participation in the sub-study, and about 60% of them (244 out of 408 individuals) agreed to participate. Subsequently, about one-fifth of these individuals (53/ 244, 21.7%) did not have an examination appointment: five individuals were unreachable; 19 individuals did not attend the agreed appointment; and with 29 individuals, an appointment arrangement was not successful or an appointment offer was declined. Data from one individual could not be included in the main analysis, because the gold standard SPPB could not be reliably conducted due to blindness of the individual. Thus, data and results from a total of 190 participants are available.

**Figure 1:**
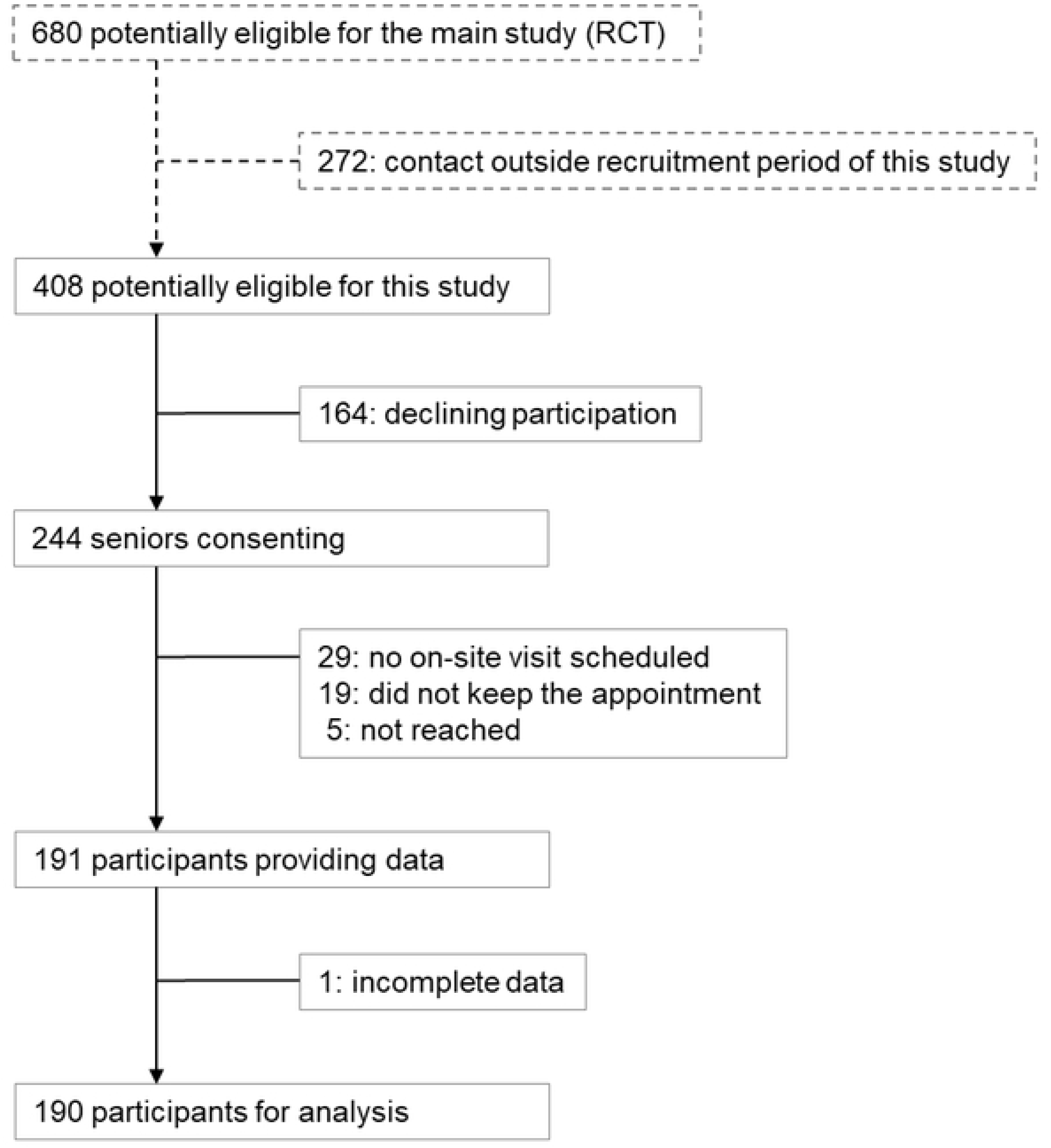
Study flow-chart

### b) Sample characteristics

The characteristics of the 190 evaluable study participants are shown in tables 1 to 3. More women than men have been recruited (143 women, 75.3%). The average age was 80.0 years ± 5.5 years (standard deviation), with women being on average about two years younger than men (79.5 versus 81.5 years, p = 0.04). In the telephone interview, half of the participants reported having fallen in the past year, a fifth reported two or more falls.

**Table 1:** Characteristics of study participants

| Characteristic | Frequency<br>(*n* = 190) | Proportion<br>(%) |
| --- | --- | --- |
| Female sex | 143 | 75.3 |
| Age (years) |  |  |
| ≥ 70 to < 75 | 38 | 20.0 |
| ≥ 75 to < 80 | 54 | 28.4 |
| ≥ 80 to < 85 | 60 | 31.6 |
| ≥ 85 to < 90 | 28 | 14.7 |
| ≥ 90 | 10 | 5.3 |
| Higher educational level (at least ten years of education) | 105 | 55.3 |
| Body-mass-index (kg/m<sup>2</sup>) |  |  |
| < 22 | 28 | 14.7 |
| ≥ 22 to < 30 | 115 | 60.6 |
| ≥ 30 | 47 | 24.7 |
| Smoking habits |  |  |
| current smoking | 10 | 5.3 |
| former smoking | 109 | 57.4 |
| German care grade | 40 | 21.1 |
| Current use of a walker | 39 | 20.5 |
| Falls (self-report) |  |  |
| at least one fall | 96 | 50.5 |
| frequent falls (≥ 2 falls) | 40 | 21.1 |

Abbreviations: kg: kilogram, m: meter.

**Table 2:** Comorbidities analogous to the Charlson Comorbidity Index

| **Comorbidity** | **Frequency**<br>(*n* = 190) | **Proportion**<br>(%) |
| --- | --- | --- |
| CCI score |  |  |
| 0 pts | 62 | 32.6 |
| 1-2 pts | 70 | 36.8 |
| 3-4 pts | 36 | 18.9 |
| ≥ 5 pts | 22 | 11.6 |
| Myocardial infarction | 12 | 6.3 |
| Chronic heart failure | 45 | 23.7 |
| Peripheral arterial disease | 17 | 8.9 |
| Stroke history | 15 | 7.9 |
| Hemiplegia | 4 | 2.1 |
| Dementia | 10 | 5.3 |
| Chronic obstructive lung disease | 40 | 21.1 |
| Rheumatic disease | 14 | 7.4 |
| Peptic ulcer disease | 22 | 11.5 |
| Diabetes mellitus | 32 | 16.8 |
| Moderate to severe kidney failure | 28 | 14.7 |
| Cancer including leukemia | 50 | 26.3 |
| Moderate to severe liver disease | 28 | 14.7 |
| HIV infection | 1 | 0.5 |

**Abbreviations:** CCI: Charlson Comorbidity Index, pts: points, HIV: Human Immunodeficiency Virus.

**Table 3:**
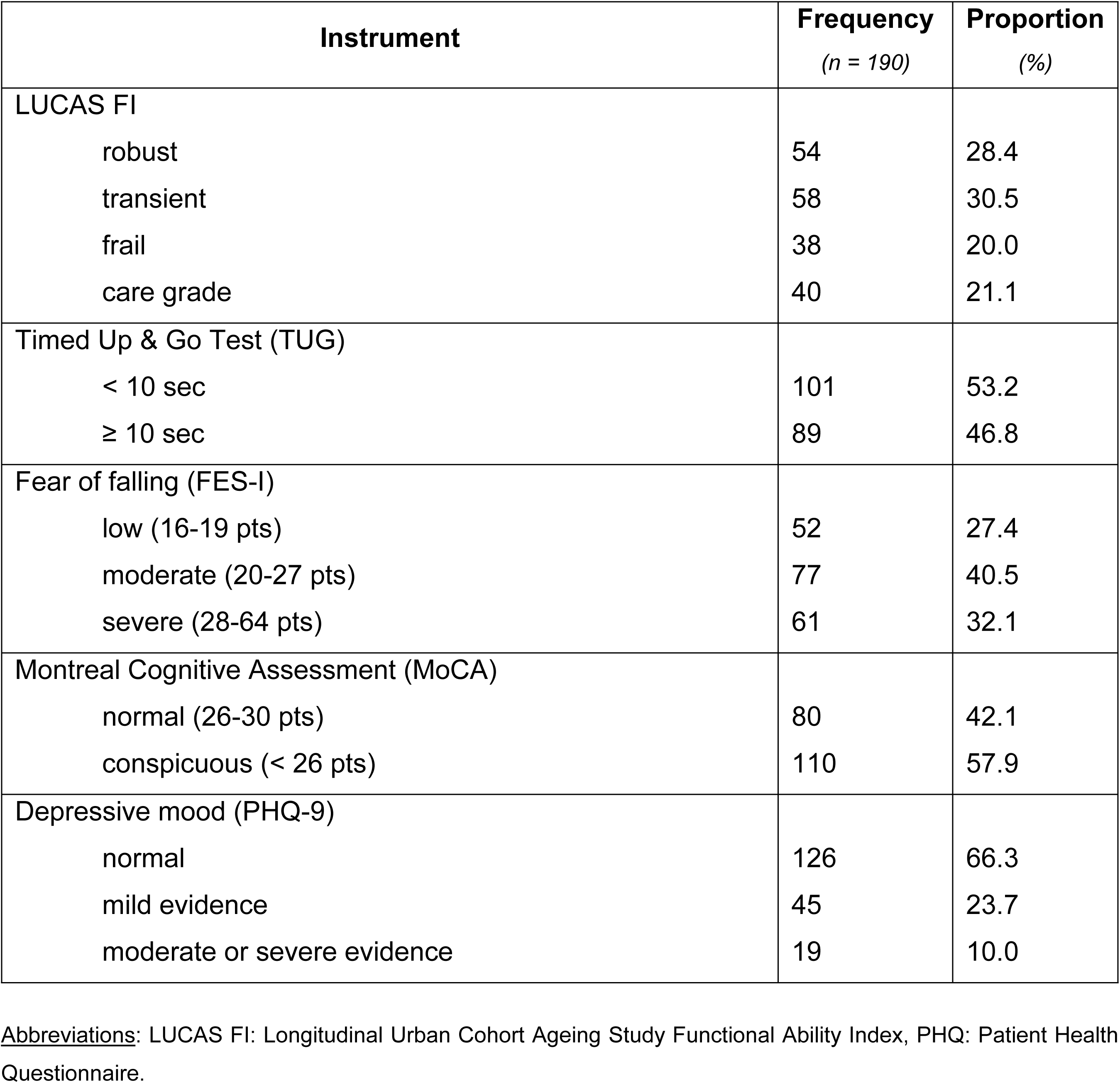
Functional status of study participants

An overview of comorbidities captured analogously to the Charlson Comorbidity Index (CCI) is given in table 2. According to the CCI score, about 30% suffered from advanced comorbidity (CCI ≥ 3 points). The most common diseases covered by the CCI were chronic heart failure (23.7%), chronic lung diseases (21.1%), and diabetes mellitus (16.8%). A positive cancer history was also common in about a quarter of cases. Among other self-reported diseases, high blood pressure (70.0%), osteoarthritis (68.4%), vertigo (60.5%), and lipid metabolism disorders (53.2%) were most frequently mentioned. Sensory impairments (hearing and vision impairment, both around 40%) were also common. Further details can be found in the supplements (tables S2 and S3).

Table 3 summarizes the key results of the geriatric assessment. According to the LUCAS FI, about 20% of the still independently living participants have advanced functional impairment (frail group). Additionally, another 40 individuals (21.1%) have a German care grade. Together, both groups thus make up more than 40% of individuals with advanced functional impairment. More than four out of ten participants show a slow Timed Up & Go Test (time ≥ 10 sec., 46.8%). In the majority of cases, increased fear of falling is present. Over half of the participants are conspicuous in screening for cognitive impairments, about a third in depression screening.

A comparison of selected characteristics according to falls risk as assessed by SIQ is depicted in table 4. Overall, participants at falls risk reported more comorbidities and worser self-perceived health, and they appeared to be more affected by functional impairment in various functional domains. A comparison of single comorbidities can be found in the supplements (table S3).

**Table 4:** Selected characteristics according to falls risk assessed by the Stay Independent Questionnaire

| Characteristic | SIQ positive<br>(n = 148) |  | SIQ negative<br>(n = 42) |  | total<br>(n = 190) |  | p-value |
| --- | --- | --- | --- | --- | --- | --- | --- |
|  | n | % | n | % | n | % |  |
| Female sex | 107 | 72.3 | 36 | 85.7 | 143 | 75.3 | > 0.05 |
| Age ≥ 80 years | 81 | 54.7 | 17 | 40.5 | 98 | 51.6 | > 0.05 |
| BMI ≥ 30 kg/m<sup>2</sup> | 40 | 27.0 | 7 | 16.7 | 47 | 24.7 | > 0.05 |
| Smoking (current or former) | 88 | 59.5 | 31 | 73.8 | 119 | 62.6 | > 0.05 |
| Higher education | 46 | 31.1 | 13 | 31.0 | 59 | 31.1 | > 0.05 |
| Low self-perceived health | 87 | 58.8 | 6 | 14.3 | 93 | 49.0 | <0.001 |
| CCI > 2 pt. | 52 | 35.1 | 6 | 14.3 | 58 | 30.5 | < 0.05 |
| German Care grade | 38 | 25.7 | 2 | 4.8 | 40 | 21.1 | 0.001 |
| TUG ≥ 10 sec | 88 | 59.5 | 1 | 2.4 | 89 | 46.8 | <0.001 |
| MoCA < 26 pt. | 89 | 60.1 | 21 | 50.0 | 110 | 57.9 | > 0.05 |
| PHQ-9 category depression | 59 | 39.9 | 5 | 11.9 | 64 | 33.7 | <0.001 |
| SARC-F ≥ 4 pt. | 74 | 50.0 | 2 | 4.8 | 76 | 40.0 | <0.001 |
| FES-I ≥ 14 pt. | 60 | 40.5 | 1 | 2.4 | 61 | 32.1 | <0.001 |
| SPPB < 10 pt. | 77 | 52.0 | 4 | 9.5 | 81 | 42.6 | <0.001 |

**Abbreviations:** CCI: Charlson Comorbidity Index, FES-I: Falls Efficacy Scale International, kg: kilogram, m: meter, MoCA: Montreal Cognitive Assessment, PHQ: Patient Health Questionnaire, pt: points, SARC-F: Strength, Assistance with walking, Rising from a chair, Climbing stairs, and Falls, sec: seconds, SIQ: Stay Independent Questionnaire, SPPB: Short Physical Performance Battery, TUG: Timed Up & Go Test.

### c) Stay Independent Questionnaire as a diagnostic test

According to the translated version of the SIQ, applied by telephone interview, 148 of the 190 participants (77.9%) have been classified as being at increased falls risk. The results according to SIQ were unambiguous, according to the CDC recommendations for its use. The SPPB examination found impairments in standing and walking balance in 81 of 190 participants (42.6%). In the walking part of the SPPB alone, with measurement of walking speed, 37 participants (19.5%) were pathological, i.e., with a walking speed < 0.8 meters per second. There were no complications, hazardous situations, or falls during the SPPB performance. The SPPB evaluation was also unambiguous according to the existing recommendations for performing and evaluating the SPPB.

The median time span between conducting the telephone interview with SIQ and the examination appointment with SPPB performance was on average 46 days (range: 9 to 211 days). For six participants, the time span exceeded 90 days. The reported results, particularly regarding the test quality of SIQ, do not change when these six cases are excluded from the analysis (data not shown).

Figure 2 shows the distribution of the total score of SIQ without special weighting of the question about a fall in the past year. Lower values in SIQ indicate lower fall risk. Figure 3 shows the distribution of the score values of the SPPB. In SPPB, lower score values indicate increasing impairment of standing and walking balance.

**Figure 2:**
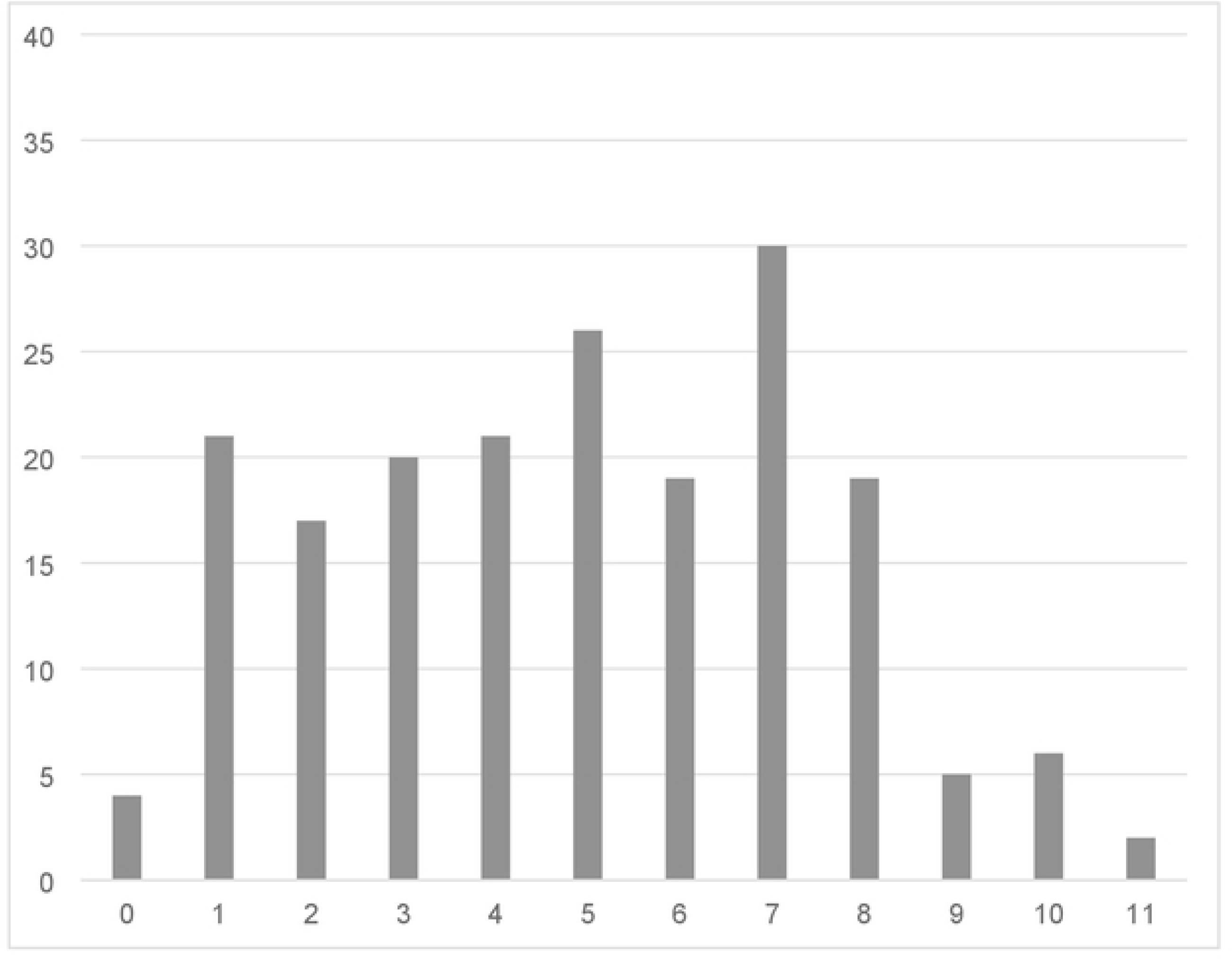
Distribution of the Stay Independent Qestionnaire (SIQ) total score. Legend: X axis: SIQ total score values (minimum 0 points), with higher values indicating (increasing) risk of falling; Y axis: absolute number of cases. The distribution of the total score of SIQ is depicted without special weighting of the question about a fall in the past year, i. e. with each item counting one point. Lower values in SIQ indicate lower fall risk.

**Figure 3:**
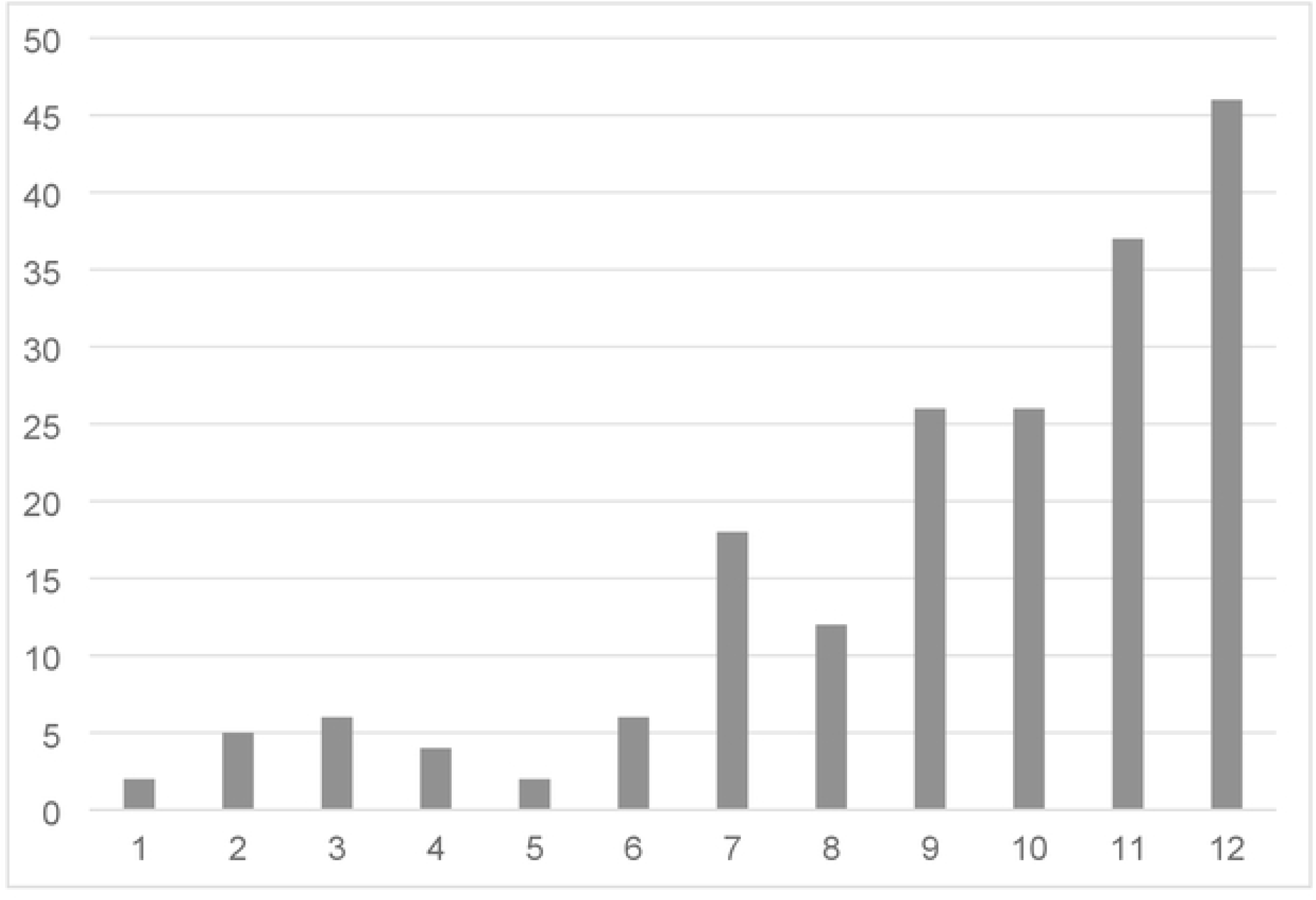
Distribution of the Short Physical Performance Battery (SPPB) total score. Legend: X axis: SPPB total score values (maximum 12 points), with lower values indicating (increasing) impairments in standing and/or walking balance; Y axis: absolute number of cases.

In figure 4, the sample distribution according to the diagnostic test SIQ and the gold standard SPPB is depicted as a two-by-two contingency table. Measured against the gold standard, SIQ correctly identifies 77 individuals as true positive and 38 individuals as true negative. Of 148 SIQ positive individuals, 71 are unimpaired in standing and walking balance according to SPPB, i.e., false positive. Of the 42 test-negative individuals, four are actually impaired, i.e. false negative.

**Figure 4:**
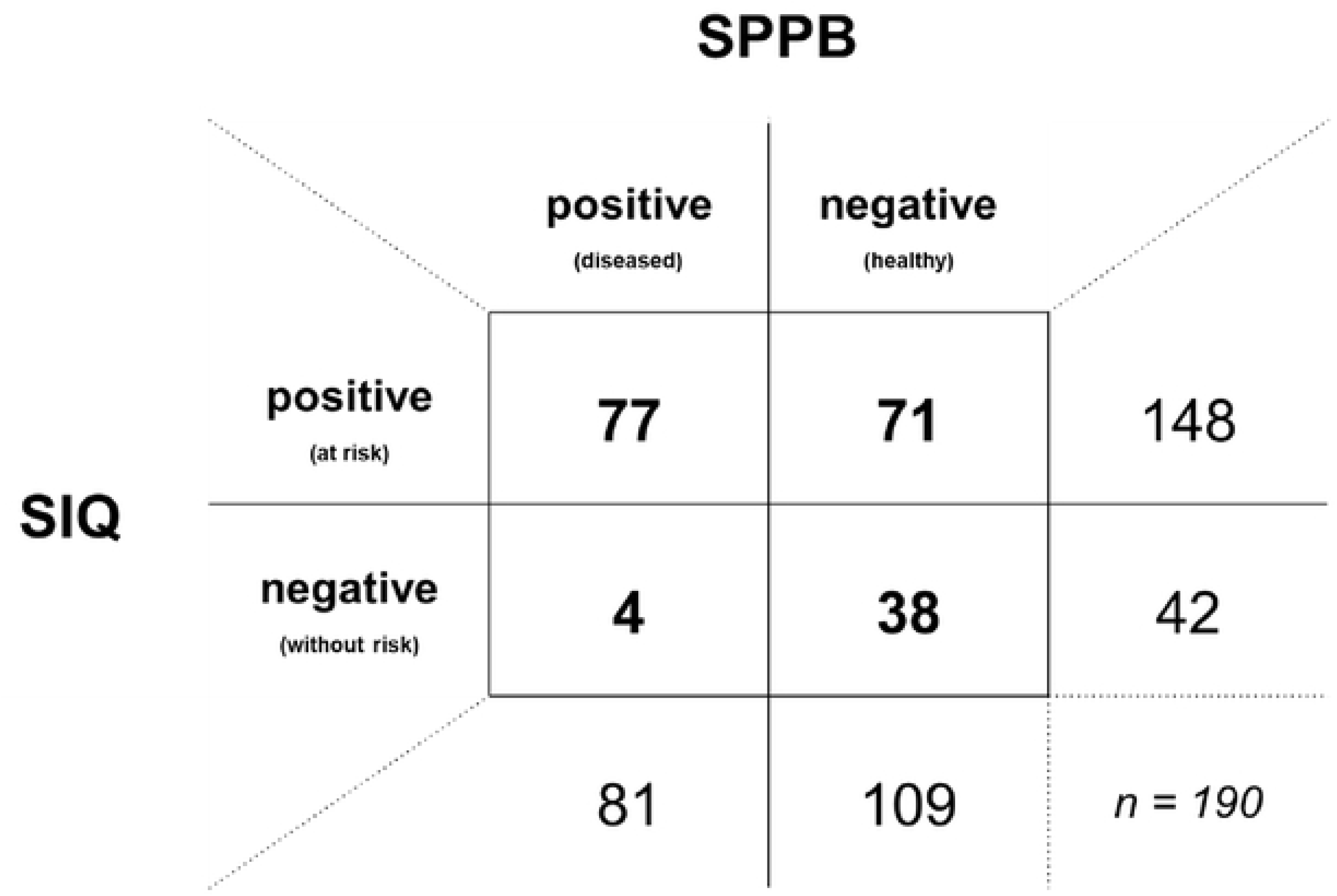
Contingency table of Stay Independent Questionnaire (SIQ) and Short Physical Performance Battery (SPPB) Legend: SPPB was used as the gold standard, with a total SPPB score < 10 points defining disease (i.e.. impairment in standing and/or walking balance);diagnostic test Stay Independent Questionnaire (SIQ), defining risk of falling with a total score of ≥ 4 points.

Table 5 lists the statistical measures of test performance. Accordingly, SIQ as a diagnostic test has a measured sensitivity of 95.1%, higher than assumed (75%). At the same time, the specificity is low at 34.9%. As assumed for the sample size calculation, the 95% CI for both sensitivity and specificity are < 10%. Corresponding to sensitivity and specificity, a slightly increased positive likelihood ratio (LR+ = 1.46) and a significantly decreased negative likelihood ratio (LR-= 0.14) are found.

**Table 5:** Performance measures of the Stay Independent Questionnaire (SIQ)

| Performance measure |  | Estimate | 95% CI |
| --- | --- | --- | --- |
| *Sn* | Sensitivity | 95.1 | [ 87.8 ; 98.6 ] |
| *Sp* | Specificity | 34.9 | [ 26.0 ; 44.6 ] |
| *PPV* | Positive predictive value | 52.0 | [ 43.6 ; 60.3 ] |
| *NPV* | Negative predictive value | 90.5 | [ 77.4 ; 97.3 ] |
| *LR+* | Positive likelihood ratio | 1.46 | [ 1.26 ; 1.69 ] |
| *LR-* | Negative likelihood ratio | 0.14 | [ 0.05 ; 0.38 ] |

Abbreviation: 95% CI: 95% Confidence Interval.

The relationship between sensitivity and specificity emerges graphically from the receiver operating characteristic (ROC) curve, which is shown in figures 5a and b. In unadjusted analysis (figure 5a), an area under the curve (AUC) of 65% indicates a rather low to moderate overall performance of SIQ. After adjustment for age and sex, performed with calculated probabilities from a logistic regression model, the test performance improves somewhat, to an AUC of 71% (figure 5b).

**Figure 5a:**
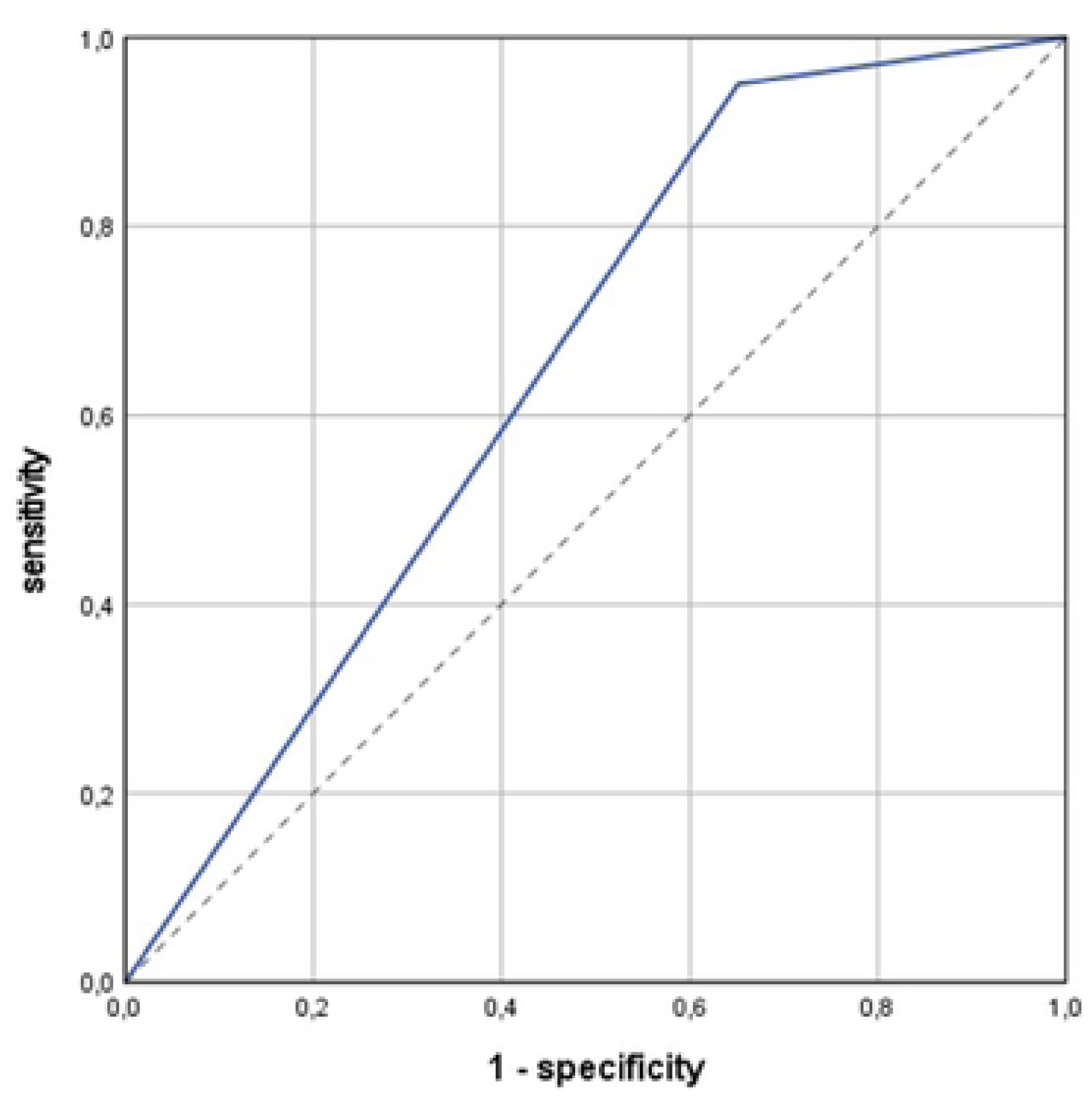
Global test performance of Stay Independent Questionnaire in unadjusted ROC analysis. Legend: Receiver operating characteric (ROC) curve analysis; target condition: impaired standing and walking balance, as assessed by Short Physical Performance Battery (SPPB score < 10 points); diagnostic test: Stay Independent Questionnaire (SIQ) defined risk of falls (SIQ score ≥ 4 points); blue line: ROC curve; dotted line: reference line; area under the curve (AUC): 0.65 [ 0.57 ; 0.73); unadjusted analysis.

**Figure 5b:**
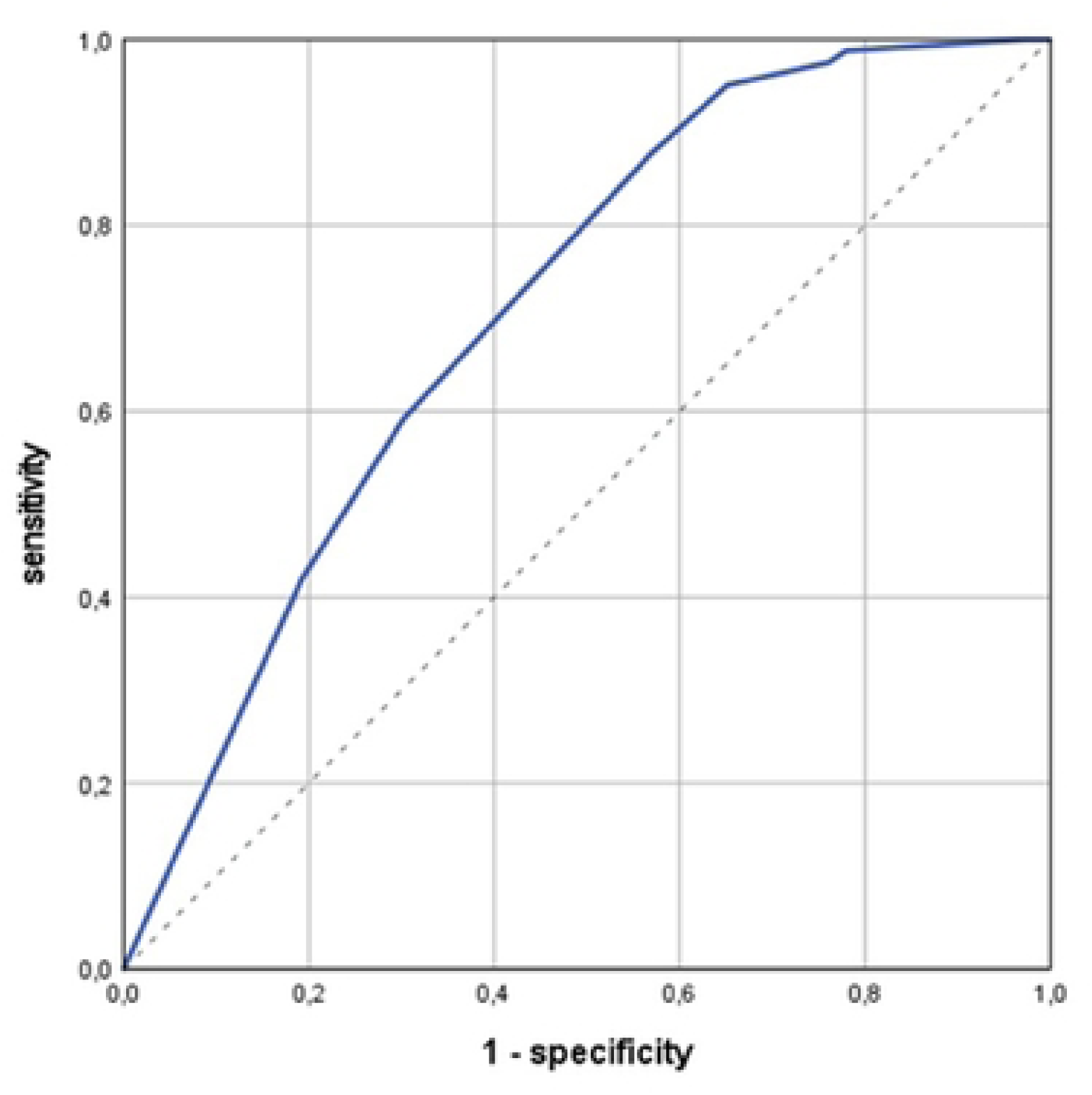
Global test performance of Stay Independent Questionnaire in ROC analysis adjusted for age and sex. Legend: Receiver operating characteric (ROC) curve analysis; target condition: impaired standing and walking balance, as assessed by Short Physical Performance Battery (SPPB score < 10 points); diagnostic test: Stay Independent Questionnaire (SIQ) defined risk of falls (SIQ score ≥ 4 points); blue line: ROC curve; dotted line: reference line; area under the curve (AUC): 0.71 [ 0.64; 0.78); adjustment for age and sex.

## Discussion

The project described here has translated the Stay Independent Questionnaire used in the American STEADI prevention program (20) for assessing fall risk in seniors into German and validated it against a standard mobility assessment test, SPPB (31). This provides another tool for use in German-speaking countries. The advantage of SIQ primarily lies in its ease of use. It demands answering just twelve relatively straightforward questions with a yes or no. The Fall Risk Check, a tool from our research group, covers 13 areas. However, it requires checking and making between two and seven individual statements per area, which is more cumbersome than SIQ (13). The Obrist questionnaire even includes 29 predictors for fall risk and has a total of 36 questions (18). Thus, both previously available German-language tools are more comprehensive and cumbersome than the SIQ questionnaire tested here. In contrast to SIQ, both would also be less suitable for use in a telephone interview.

The main feature of SIQ, according to the data presented here, is its high sensitivity (∼95%). This qualifies SIQ as a simple test that reliably identifies seniors with unimpaired standing and walking balance, excluding these individuals from further, presumably unnecessary examinations. The primary purpose of SIQ in the INES project was achieved, namely to exclude insured persons with no or only minor balance impairments and, consequently, presumably no or only minor fall risk from participation in the main study. A drawback of SIQ is the low specificity measured at about 35%, which means that many individuals without objectively measurable balance impairments are still among those tested positive.

Comparing SIQ’s test performance with the two German-language instruments is hardly feasible. For the Fall Risk Check, there is no validation study yet that tests the instrument against a mobility assessment or against the endpoint of falls over time (19). Obrist et al. validate their online questionnaire against incident falls over six months with monthly follow-up reporting, but do not report sensitivity or specificity or raw data. Only the overall quality from ROC analysis is reported, with an AUC of 0.67 (18). Comparing the two study populations shows that participants in our study were significantly older (nearly ten years difference) than those in the Obrist study, reported falls more frequently in the past twelve months (about 50% versus about 20%), and generally seem to suffer from significantly more advanced morbidity.

Regarding other instruments, the data presented on SIQ’s test performance appear comparable. Most publications report AUC values as an expression of the overall test performance. AUC values between 68% and 73% are described for most tools (14,16,17). Alessi et al. report a sensitivity of 94% and a specificity of 51% for GPSS, the latter slightly higher but comparable to the values for SIQ (12). Results for FROP-COM with a sensitivity of 93% and a specificity of 30%, measured at a score threshold of 2 points, are also comparable (14). A strength of most mentioned studies is that they often validate against newly occurring, i.e. incident falls during follow-up (12,14,16,17). While other studies use a twelve-month follow-up period, Hirase’s study (16) only follows up for three months, limiting comparability. Unfortunately, it was not possible to conduct a follow-up examination of participants within our project. While incident falls could generally be recorded among participants of the main study, who continue to be followed in the INES project, those testing negative in SIQ, who were excluded from the main study, cannot be questioned and observed any further. We, therefore, opted for a cross-sectional assessment of mobility data and validated the questionnaire against SPPB.

Choosing a gold standard for mobility assessment is still challenging. Current evidence does not clearly indicate which assessment is best suited for detecting mobility impairment or identifying a risk of falling. In the World Fall Guidelines, walking speed, alternatively the Timed Up & Go Test, is preferred over other assessments (1) for the group at intermediate fall risk. Most recent systematic reviews also recommend walking speed for walking tests, alternatively Timed Up & Go Test, Berg Balance Scale, Performance Oriented Mobility Assessment, or Tandem and Semi-Tandem Stance for balance testing (52–56). The Chair Rise Test and its modifications are also frequently mentioned (52,53). As already described, we have chosen SPPB as the gold standard. The recommendation of the World Fall Guidelines is deemed unsuitable for this study’s purpose, as we needed a gold standard for all participants, not just for a specific risk group. In communicating about standing and walking ability to test persons, a multi-part assessment like SPPB has advantages over other instruments, which was important for us in the study. Extensive application experience in our study centre has also been an argument for SPPB. Finally, walking speed, Tandem and Semi-Tandem Stance, and Chair Rise Test are included in the three parts of SPPB, so these parameters can be used for further analysis, albeit secondarily.

Pure self-report instruments like SIQ necessarily have limitations. Interestingly, instruments that combine self-reports from test persons, demographic data, and data from assessment or performance tests do not necessarily perform better in terms of test performance criteria. AUC values between 64% and 71% are reported for many instruments and models (57–66). Data from the modified FROP-COM deviate somewhat, with an AUC of 0.79 (67). In this tool, age over 80 years, female sex, two or more falls, and a tendency toward depression (according to SCL90), hand strength, and postural sway while standing are combined (67). However, neither this nor any other model has yet established itself as a standard. In a comprehensive systematic review using rigorous criteria for study quality, Gade et al. (68) recently concluded that none of the existing proposals for predicting falls among independently living, community-dwelling seniors is suitable for standard application.

The study presented here has limitations that must be considered when interpreting the results. First, it must be emphasized that SIQ used in STEADI is intended as a self-completion tool and is also used as such. We used the German version of SIQ in a telephone interview. Whether the deviations between self-completion and telephone interview are significant is unclear. However, we cannot quantify such a deviation. Therefore, follow-up studies using SIQ as a self-completion tool should follow. Secondly, it must be noted that the preselection of insured persons, at least at the beginning of recruitment, also included coded ICD-10 codes and thus diseases associated with an increased risk of falls. In our view, this does not necessarily affect measures such as sensitivity and specificity, especially since the disease-based preselection has been abandoned in the second half of the recruitment. But certainly, not all characteristics of the participants presented here can automatically be transferred to all seniors living independently. Finally, it should be mentioned that the sample of insured persons in Hamburg examined in our subproject is not a random sample of the entire INES study population. This means that the characteristics of those individuals included in the INES main study as at-risk of falling from all three regions may well differ from the characteristics of the study population in Hamburg that we examined.

## Conclusions

The German translation of the SIQ questionnaire is capable of identifying community-dwelling older people with unimpaired standing and walking balance, as measured by SPPB.

Thus, SIQ appears to fulfil the main requirement of a screening test for falls risk, i.e., to exclude unimpaired, balance-healthy seniors with presumably low risk.

The recruited group of seniors is characterized by a moderate disease burden, clear functional limitations, and an overall at least moderately increased fall risk. This might also be expected for the entire study population of the ongoing RCT.

## Data Availability

Data from this study are not publicly available due to legal issues, especially German data protection regulations. The Ethics Committee of the Hamburg Medical Association did only allow data collection, processing and storage for this particular project and research question. Furthermore, participants have not been informed about a secondary use of data by other researchers, and hence did not give consent to this. A further use of our data would demand a new application at the Ethics Committee of the Hamburg Medical Association. We will be ready to assist other researchers with interest in using our data in a new application at the responsible ethics committee, and we will be happy to provide data in case of a new positive vote.

## Acknowledgements

We gratefully acknowledge Ms. S. Stasch for excellent secretarial work and support, and Ms. B. Ruß-Thiel and Mr. T. Schaknat, ife Gesundheits-GmbH, Nehmten, Germany, for their cooperation and for organising and performing the telephone interviews. Our sincere thank goes to all study participants and their relatives interested in our work and supporting this study.

## References

1. Montero-Odasso M, van der Velde N, Martin FC, Petrovic M, Tan MP, Ryg J, et al. World guidelines for falls prevention and management for older adults: a global initiative. Age Ageing. 2. September 2022;51(9).

2. Moreland B, Kakara R, Henry A. Trends in Nonfatal Falls and Fall-Related Injuries Among Adults Aged ≥65 Years - United States, 2012-2018. MMWR Morb Mortal Wkly Rep. 10. Juli 2020;69(27):875–81.

3. Lu H, Dong XX, Li DL, Wu Q, Nie XY, Xu Y, et al. Prevalent falls, fall frequencies and health-related quality of life among community-dwelling older Chinese adults. Qual Life Res. November 2023;32(11):3279–89.

4. Miri S, Norasteh AA. Fear of falling, quality of life, and daily functional activity of elderly women with and without a history of falling: a cross-sectional study. Ann Med Surg (Lond). Mai 2024;86(5):2619–25.

5. Montero-Odasso M, van der Velde N, Alexander NB, Becker C, Blain H, Camicioli R, et al. New horizons in falls prevention and management for older adults: a global initiative. Age Ageing. 11. September 2021;50(5):1499–507.

6. DeStatis (Statistisches Bundesamt). Statistiken zu Diagnosen von Krankenhauspatienten [Internet]. online; 2024. https://www-genesis.destatis.de/genesis/online?operation=statistic&levelindex=0&levelid=1718051471116&code=23131#abreadcrumb

7. Sebastiani C, Wong JYX, Litt A, Loewen J, Reece K, Conlin N, et al. Mapping sex and gender differences in falls among older adults: A scoping review. J Am Geriatr Soc. März 2024;72(3):903–15.

8. Burns ER, Stevens JA, Lee R. The direct costs of fatal and non-fatal falls among older adults - United States. J Safety Res. September 2016;58:99–103.

9. Florence CS, Bergen G, Atherly A, Burns E, Stevens J, Drake C. Medical Costs of Fatal and Nonfatal Falls in Older Adults. J Am Geriatr Soc. April 2018;66(4):693–8.

10. Ou W, Zhang Q, He J, Shao X, Yang Y, Wang X. Hospitalization costs of injury in elderly population in China: a quantile regression analysis. BMC Geriatr. 14. März 2023;23(1):143.

11. Hoffman GJ, Liu H, Alexander NB, Tinetti M, Braun TM, Min LC. Posthospital Fall Injuries and 30-Day Readmissions in Adults 65 Years and Older. JAMA Netw Open. 3. Mai 2019;2(5):e194276.

12. Alessi CA, Josephson KR, Harker JO, Pietruszka FM, Hoyl MT, Rubenstein LZ. The yield, reliability, and validity of a postal survey for screening community-dwelling older people. J Am Geriatr Soc. Februar 2003;51(2):194–202.

13. Anders J, Dapp U, Laub S, von Renteln-Kruse W, Juhl K. [Screening of fall risk in frail, but still independently living senior citizens]. Z Gerontol Geriatr. August 2006;39(4):268–76.

14. Russell MA, Hill KD, Blackberry I, Day LM, Dharmage SC. The reliability and predictive accuracy of the falls risk for older people in the community assessment (FROP-Com) tool. Age Ageing. November 2008;37(6):634–9.

15. Rubenstein LZ, Vivrette R, Harker JO, Stevens JA, Kramer BJ. Validating an evidence-based, self-rated fall risk questionnaire (FRQ) for older adults. J Safety Res. Dezember 2011;42(6):493–9.

16. Hirase T, Inokuchi S, Matsusaka N, Nakahara K, Okita M. A modified fall risk assessment tool that is specific to physical function predicts falls in community-dwelling elderly people. J Geriatr Phys Ther. Dezember 2014;37(4):159–65.

17. Gadkaree SK, Sun DQ, Huang J, Varadhan R, Agrawal Y. Comparison of simple vs. performance-based fall prediction models: data from the National Health and Aging Trends Study. Gerontol Geriatr Med. Dezember 2015;1:2333721415584850.

18. Obrist S, Rogan S, Hilfiker R. Development and Evaluation of an Online Fall-Risk Questionnaire for Nonfrail Community-Dwelling Elderly Persons: A Pilot Study. Curr Gerontol Geriatr Res. 2016;2016:1520932.

19. Anders J, Dapp U, Laub S, von Renteln-Kruse W. [Impact of fall risk and fear of falling on mobility of independently living senior citizens transitioning to frailty: screening results concerning fall prevention in the community]. Z Gerontol Geriatr. August 2007;40(4):255–67.

20. Stevens JA. The STEADI Tool Kit: A Fall Prevention Resource for Health Care Providers. IHS Prim Care Provid. September 2013;39(9):162–6.

21. Lafontant K, Blount A, Suarez JRM, Fukuda DH, Stout JR, Trahan EM, et al. Comparing Sensitivity, Specificity, and Accuracy of Fall Risk Assessments in Community-Dwelling Older Adults. Clin Interv Aging. 2024;19:581–8.

22. Loonlawong S, Limroongreungrat W, Rattananupong T, Kittipimpanon K, Saisanan Na Ayudhaya W, Jiamjarasrangsi W. Predictive validity of the Stopping Elderly Accidents, Deaths & Injuries (STEADI) program fall risk screening algorithms among community-dwelling Thai elderly. BMC Med. 14. März 2022;20(1):78.

23. Mielenz TJ, Kannoth S, Jia H, Pullyblank K, Sorensen J, Estabrooks P, et al. Evaluating a Two-Level vs. Three-Level Fall Risk Screening Algorithm for Predicting Falls Among Older Adults. Front Public Health. 2020;8:373.

24. Nithman RW, Vincenzo JL. How steady is the STEADI? Inferential analysis of the CDC fall risk toolkit. Arch Gerontol Geriatr. August 2019;83:185–94.

25. Camp K, Murphy S, Pate B. Integrating Fall Prevention Strategies into EMS Services to Reduce Falls and Associated Healthcare Costs for Older Adults. Clin Interv Aging. 2024;19:561–9.

26. McGee S. Evidence-Based Physical Diagnosis. 1. Auflage. Saunders; 2001.

27. Bundesinstitut für Arzneimittel und Medizinprodukte. ICD-10-GM: Internationale statistische Klassifikation der Krankheiten und verwandter Gesundheitsprobleme, German Modification [Internet]. online; 2023. https://www.bfarm.de/DE/Kodiersysteme/Klassifikationen/ICD/ICD-10-GM/_node.html

28. Stevens JA, Phelan EA. Development of STEADI: a fall prevention resource for health care providers. Health Promot Pract. September 2013;14(5):706–14.

29. Center for Disease Control and Prevention. Coordinated Care Plan to Prevent Older Adults Falls [Internet]. CDC; 2021. https://www.cdc.gov/steadi/hcp/clinical-resources/outpatient-care.html

30. Penninx BW, Ferrucci L, Leveille SG, Rantanen T, Pahor M, Guralnik JM. Lower extremity performance in nondisabled older persons as a predictor of subsequent hospitalization. J Gerontol A Biol Sci Med Sci. November 2000;55(11):M691–697.

31. Guralnik JM, Simonsick EM, Ferrucci L, Glynn RJ, Berkman LF, Blazer DG, et al. A short physical performance battery assessing lower extremity function: association with self-reported disability and prediction of mortality and nursing home admission. J Gerontol. März 1994;49(2):M85–94.

32. Podsiadlo D, Richardson S. The timed „Up & Go“: a test of basic functional mobility for frail elderly persons. J Am Geriatr Soc. Februar 1991;39(2):142–8.

33. Dias N, Kempen GIJM, Todd CJ, Beyer N, Freiberger E, Piot-Ziegler C, et al. [The German version of the Falls Efficacy Scale-International Version (FES-I)]. Z Gerontol Geriatr. August 2006;39(4):297–300.

34. Kempen GIJM, Yardley L, van Haastregt JCM, Zijlstra GAR, Beyer N, Hauer K, et al. The Short FES-I: a shortened version of the falls efficacy scale-international to assess fear of falling. Age Ageing. Januar 2008;37(1):45–50.

35. Drey M, Ferrari U, Schraml M, Kemmler W, Schoene D, Franke A, et al. German Version of SARC-F: Translation, Adaption, and Validation. J Am Med Dir Assoc. Juni 2020;21(6):747–751.e1.

36. Malmstrom TK, Morley JE. SARC-F: a simple questionnaire to rapidly diagnose sarcopenia. J Am Med Dir Assoc. August 2013;14(8):531–2.

37. Hamilton GF, McDonald C, Chenier TC. Measurement of grip strength: validity and reliability of the sphygmomanometer and jamar grip dynamometer. J Orthop Sports Phys Ther. 1992;16(5):215–9.

38. Tinsley GM, Moore ML, Silva AM, Sardinha LB. Cross-sectional and longitudinal agreement between two multifrequency bioimpedance devices for resistance, reactance, and phase angle values. Eur J Clin Nutr. Juni 2020;74(6):900–11.

39. Nasreddine ZS, Phillips NA, Bédirian V, Charbonneau S, Whitehead V, Collin I, et al. The Montreal Cognitive Assessment, MoCA: a brief screening tool for mild cognitive impairment. J Am Geriatr Soc. April 2005;53(4):695–9.

40. Löwe B, Kroenke K, Herzog W, Gräfe K. Measuring depression outcome with a brief self-report instrument: sensitivity to change of the Patient Health Questionnaire (PHQ-9). J Affect Disord. Juli 2004;81(1):61–6.

41. Dapp U, Minder CE, Anders J, Golgert S, von Renteln-Kruse W. Long-term prediction of changes in health status, frailty, nursing care and mortality in community-dwelling senior citizens—results from the Longitudinal Urban Cohort Ageing Study (LUCAS). BMC Geriatr. 19. Dezember 2014;14:141.

42. Greiner W, Weijnen T, Nieuwenhuizen M, Oppe S, Badia X, Busschbach J, et al. A single European currency for EQ-5D health states. Results from a six-country study. Eur J Health Econ. September 2003;4(3):222–31.

43. Marten O, Greiner W. Feasibility properties of the EQ-5D-3L and 5L in the general population: evidence from the GP Patient Survey on the impact of age. Health Econ Rev. 20. Mai 2022;12(1):28.

44. Charlson ME, Pompei P, Ales KL, MacKenzie CR. A new method of classifying prognostic comorbidity in longitudinal studies: development and validation. J Chronic Dis. 1987;40(5):373–83.

45. Quan H, Li B, Couris CM, Fushimi K, Graham P, Hider P, et al. Updating and validating the Charlson comorbidity index and score for risk adjustment in hospital discharge abstracts using data from 6 countries. Am J Epidemiol. 15. März 2011;173(6):676–82.

46. Hajian-Tilaki K. Sample size estimation in diagnostic test studies of biomedical informatics. J Biomed Inform. April 2014;48:193–204.

47. Simel DL, Samsa GP, Matchar DB. Likelihood ratios with confidence: sample size estimation for diagnostic test studies. J Clin Epidemiol. 1991;44(8):763–70.

48. Wilson EB. Probable Inference, the Law of Succession, and Statistical Inference. Journal of the American Statistical Association. 1927;22(158):209–2012.

49. Agresti A, Coull B. Approximate is Better than „Exact“ for Interval Estimation of Binomial Proportions. The American Statistician. 1998;52(2):119–26.

50. WMA (World Medical Association). World Medical Association Declaration of Helsinki: ethical principles for medical research involving human subjects. JAMA. 27. November 2013;310(20):2191–4.

51. Bossuyt PM, Reitsma JB, Bruns DE, Gatsonis CA, Glasziou PP, Irwig L, et al. STARD 2015: an updated list of essential items for reporting diagnostic accuracy studies. BMJ. 28. Oktober 2015;351:h5527.

52. Beck Jepsen D, Robinson K, Ogliari G, Montero-Odasso M, Kamkar N, Ryg J, et al. Predicting falls in older adults: an umbrella review of instruments assessing gait, balance, and functional mobility. BMC Geriatr. 25. Juli 2022;22(1):615.

53. Lusardi MM, Fritz S, Middleton A, Allison L, Wingood M, Phillips E, et al. Determining Risk of Falls in Community Dwelling Older Adults: A Systematic Review and Meta-analysis Using Posttest Probability. J Geriatr Phys Ther. März 2017;40(1):1–36.

54. Marin-Jimenez N, Cruz-León C, Perez-Bey A, Conde-Caveda J, Grao-Cruces A, Aparicio VA, et al. Predictive Validity of Motor Fitness and Flexibility Tests in Adults and Older Adults: A Systematic Review. J Clin Med. 10. Januar 2022;11(2).

55. Meekes WM, Korevaar JC, Leemrijse CJ, van de Goor IA. Practical and validated tool to assess falls risk in the primary care setting: a systematic review. BMJ Open. 29. September 2021;11(9):e045431.

56. Waterval NFJ, Claassen CM, van der Helm FCT, van der Kruk E. Predictability of Fall Risk Assessments in Community-Dwelling Older Adults: A Scoping Review. Sensors (Basel). 6. September 2023;23(18).

57. Bongue B, Dupré C, Beauchet O, Rossat A, Fantino B, Colvez A. A screening tool with five risk factors was developed for fall-risk prediction in community-dwelling elderly. J Clin Epidemiol. Oktober 2011;64(10):1152–60.

58. Cattelani L, Palumbo P, Palmerini L, Bandinelli S, Becker C, Chesani F, et al. FRAT-up, a Web-based fall-risk assessment tool for elderly people living in the community. J Med Internet Res. 18. Februar 2015;17(2):e41.

59. Covinsky KE, Kahana E, Kahana B, Kercher K, Schumacher JG, Justice AC. History and mobility exam index to identify community-dwelling elderly persons at risk of falling. J Gerontol A Biol Sci Med Sci. April 2001;56(4):M253–259.

60. Hnizdo S, Archuleta RA, Taylor B, Kim SC. Validity and reliability of the modified John Hopkins Fall Risk Assessment Tool for elderly patients in home health care. Geriatr Nurs. Oktober 2013;34(5):423–7.

61. Palumbo P, Palmerini L, Bandinelli S, Chiari L. Fall Risk Assessment Tools for Elderly Living in the Community: Can We Do Better? PLoS One. 2015;10(12):e0146247.

62. Peeters GMEEG, Pluijm SMF, van Schoor NM, Elders PJM, Bouter LM, Lips P. Validation of the LASA fall risk profile for recurrent falling in older recent fallers. J Clin Epidemiol. November 2010;63(11):1242–8.

63. Pluijm SMF, Smit JH, Tromp EAM, Stel VS, Deeg DJH, Bouter LM, et al. A risk profile for identifying community-dwelling elderly with a high risk of recurrent falling: results of a 3-year prospective study. Osteoporos Int. 2006;17(3):417–25.

64. Renfro MO, Fehrer S. Multifactorial screening for fall risk in community-dwelling older adults in the primary care office: development of the fall risk assessment & screening tool. J Geriatr Phys Ther. Dezember 2011;34(4):174–83.

65. Tiedemann A, Lord SR, Sherrington C. The development and validation of a brief performance-based fall risk assessment tool for use in primary care. J Gerontol A Biol Sci Med Sci. August 2010;65(8):896–903.

66. Tromp AM, Pluijm SM, Smit JH, Deeg DJ, Bouter LM, Lips P. Fall-risk screening test: a prospective study on predictors for falls in community-dwelling elderly. J Clin Epidemiol. August 2001;54(8):837–44.

67. Stalenhoef PA, Diederiks JPM, Knottnerus JA, Kester ADM, Crebolder HFJM. A risk model for the prediction of recurrent falls in community-dwelling elderly: a prospective cohort study. J Clin Epidemiol. November 2002;55(11):1088–94.

68. Gade GV, Jørgensen MG, Ryg J, Riis J, Thomsen K, Masud T, et al. Predicting falls in community-dwelling older adults: a systematic review of prognostic models. BMJ Open. 4. Mai 2021;11(5):e044170.

